# Excess HIV infections and costs associated with reductions in HIV prevention services in the United States: Projection using real-world data

**DOI:** 10.1101/2025.04.17.25324904

**Authors:** Patrick S Sullivan, Kristin M Wall, Marta Juhasz, Greg Millett, Jeffrey Crowley, Chris Beyrer, Stephanie DuBose, Kamaria Brisco, Gordon Le, Kenneth Mayer

## Abstract

**Importance:** Pre-exposure prophylaxis (PrEP) is a proven effective intervention to reduce risk for HIV infection, but changes in policies that lead to increased out of pocket PrEP costs or that decrease access to convenient PrEP locations could reduce PrEP coverage, resulting in excess HIV infections and costs.

**Objective:** To estimate the impacts of federal policy changes on PrEP coverage, new HIV infections and costs associated with new HIV infections

**Design:** Estimation of excess HIV infections under different policy impacts were conducted using parameters from a previously published ecological model of the relationship between PrEP coverage and new HIV infections. Costs were estimated for the treatment of infections not averted under different scenarios.

**Setting:** United States

**Participants:** There was no individual participation in research activities; population-based data sources were used to describe the population-level PrEP use and new diagnoses under different hypothetical changes in PrEP coverage.

**Exposures:** Percent of people with indications for PrEP who are taking PrEP

**Main Outcome and Measures:** Estimated change in new HIV infections under different assumptions of change in PrEP coverage; costs of treatment for avoidable HIV infections and net costs of avoidable infections after accounting for costs of PrEP medications.

**Results:** Even modest reductions in PrEP coverage would result in thousands of avoidable HIV infections. An absolute 3.3% annual reduction in PrEP coverage over the next decade would result in 8,618 avoidable HIV infections, with lifetime medical costs of over $3.6 billion (discounted) for treatment of the unaverted HIV infections.

**Conclusions and Relevance:** Changes in policy that reduce PrEP uptake would result in avoidable HIV infections and increased costs for HIV treatment. Maintaining policies and programs that support PrEP uptake offers benefits for health and is estimated to result in net cost savings.

**Key points:** *Question:* What are the likely impacts on HIV transmissions and healthcare costs if policy changes result in decreased PrEP utilization in the United States?

*Findings:* Under assumptions of even modest reductions in PrEP use, we estimated thousands of HIV infections would fail to be averted over the next decade, and billions of dollars of additional treatment costs would accrue to the healthcare system.

*Results of the study:* We used historical descriptive data on the US HIV epidemic to quantify the relationship between PrEP coverage and trends in HIV diagnoses and to estimate future trends in HIV infections if PrEP coverage were to be rolled back. If PrEP use declines modestly – about 3% annually – we estimate that 8,618 new infections would fail to be averted in a decade because of lowered PrEP uptake, and the estimated lifetime medical costs of these unaverted infections would be $3.6 billion (discounted) and $9.3 billion (undiscounted).

*Meaning:* Changes in healthcare priorities and policies, especially those that increase out of pocket costs of PrEP or reduce the convenience of engaging in PrEP care, risk rolling back our progress in ending the HIV epidemic, accruing avertable HIV infections, and incurring increased costs for medical care of people whose HIV infections were avoidable.

---

Antiretroviral pre-exposure prophylaxis (PrEP) to prevent HIV acquisition is a mainstay of HIV epidemic control in the United States.^1^ PrEP, when taken as directed, reduces the risk of acquiring HIV by up to 99%.^2^ However, progress to achieve high levels of PrEP use by people who might benefit from it has been slow. In 2022, only 36% of those with PrEP indications were taking it.^3,4^ PrEP uptake is driven by multiple factors, including perceived risk of HIV infection^5^, availability of low-cost or no-cost PrEP and access to associated PrEP services^6^, proximity of knowledgeable PrEP providers^7–9^, and perceived efficacy of PrEP^5,10^.

Evidence from clinical trials documents the efficacy of PrEP for reducing HIV infections in people who take it^2^, and in ecological studies, states with high PrEP coverage among those with indications experience correspondingly lower new HIV infections.^11^ Although these associations are ecological, they are directionally consistent with the known individual efficacy of PrEP. We have recently used ecological data to demonstrate that every 5 per 100 persons with indications increase in PrEP coverage in the United States is associated with a subsequent 3.4% decline in HIV diagnosis rate, after adjusting for rates of viral suppression.^13^

Despite this demonstrated relationship between increases in PrEP coverage and decreases in new HIV diagnoses, changes in PrEP coverage over the past decade have been gradual, and there are substantial differences in the extent of PrEP coverage among US states.^13^ At the individual level, PrEP use is four times more likely in patients with health insurance^14^, and on a jurisdictional level, equitable PrEP use is substantially higher in states with Medicaid expansion or PrEP Drug Assistance Programs (PrEP-DAP), and highest in states with both.^6^ This suggests that policies related to affordability of PrEP likely influence the extent of PrEP coverage and, in turn, risks for new HIV infections in the population.

There is currently a discussion about reducing public funding for PrEP programs (i.e., increasing out of pocket costs, increased copays), even though PrEP use is cost saving when used in populations with indications.^15^ Recent cuts to staffing and programs at the Centers for Disease Control and Prevention^16^ and cuts targeting HIV-focused research funded by the National Institutes of Health are early indications that public support for HIV prevention programs and research is vulnerable. Such reductions in public financial support would likely decrease PrEP use. For example, declines in PrEP usage might occur as a result of discontinuation or reduction of Medicaid coverage or PrEP-DAP programs, or as a result of reduction or elimination of Affordable Care Act provisions that minimize out of pocket costs for PrEP and associated laboratory testing and other components of the PrEP regimen beyond the medication.^6,10^ Declines might also occur due to changes in provider behavior^17^; disinvestment in PrEP promotion campaigns; closing of publicly-supported PrEP provision sites, which would lead to increased commute times to PrEP^18^; or discontinuing provider education programs that encourage primary care providers to routinely offer PrEP to patients.^17^

To estimate the possible impact on HIV incidence and costs associated with withdrawing public supports for PrEP programs, we used previously published ecological data on the associations between PrEP coverage and HIV new diagnoses to estimate excess infections that would occur with diminished public supports for HIV prevention and PrEP (i.e., infections not averted because of decreased PrEP uptake).^11,13^ We also assessed published estimates of PrEP and treatment costs to estimate the HIV treatment costs and net costs of HIV treatment because of infections not averted.^19^

## Methods

We estimated the predicted relationship between future changes in PrEP coverage (i.e., the proportion of people with an indication for PrEP who are taking it^19^) and new HIV diagnosis by using parameters from a recent model of the real-world relationship between PrEP coverage and new diagnoses.^13^ We have previously reported methods to evaluate the relative change in HIV diagnosis rates per year for the period 2012-2022 using Poisson log-linear models from the generalized linear mixed models (GLMM) family, with the yearly new HIV diagnoses as the outcome and the log of the yearly corresponding state population denominators as an offset.^11^ We used year as an independent continuous variable to generate the Estimated Annual Percent Change (EAPC) in HIV diagnosis rates and 95% confidence intervals.^22^ The same modelling method was used to generate an Estimated Decade Percent Change (EDPC) outcome (i.e., an Estimated Cumulative Percent Change over the 10-year study period).

The associations of PrEP coverage with change in HIV diagnosis rates were assessed by including PrEP coverage as a continuous predictor in Poisson log-linear models. We reported the predicted percent change in HIV diagnosis rates for a 1 per 100 persons with indication increase, and other levels of increase in PrEP coverage as well. Analyses of the association of PrEP coverage with change in HIV rates were adjusted for contemporary state-level viral suppression rates. Models incorporated normally distributed random effects to account for between-state variation, and when year was not a fixed effect, included random effects for time to account for within-state variation. To assess whether it was appropriate to analyze the log-linear trend in HIV diagnoses without inflection points and for data visualization purposes, we used Joinpoint regression analysis.^20^ We found no inflection points and used one slope coefficient for the entire period.

For the current analysis, we evaluated the potential impact of policy changes that would result in decreasing PrEP uptake in the upcoming years. We parameterized the model using the previously reported Poisson log-linear model of the national-level ecological association between PrEP coverage and new HIV diagnosis. To present the counterfactual scenarios of changes in new HIV diagnoses in future years, we used data from the original real-world model^13^ and we reversed the sign of the negative parameter estimate (e.g., coefficient represented declines in HIV diagnoses associated with increasing PrEP use) to obtain the predicted increases in new HIV diagnosis rates associated with several hypothesized levels of decrease in PrEP coverage. We calculated the predicted percent increase in HIV diagnosis rates for a 1 unit (1 per 100 persons with indication) decrease in PrEP coverage compared to the original inverse model. We applied these estimates to three possible future scenarios of PrEP coverage. *Scenario 1* reflected an assumption that the increases in PrEP coverage observed over the past decade would be reversed over the following decade. We calculated the degree of decrease in PrEP coverage that would result in the reversal of the estimated annual percent change in our original estimation, averaged across a decade (2012-2022) and adjusted for viral suppression rates at the state level: a 3.3 per 100 average annual decrease in PrEP coverage approximated a reversal of gains from the prior decade (e.g., a 2.3% annual increase in HIV diagnosis rates). We then applied these percentages to the new HIV diagnosis rate and to the PrEP coverage, reported in 2022 as baseline values. We calculated the projected number of persons not receiving PrEP over the next decade given the reversal of prior decade gains in PrEP coverage, and the resulting number of HIV infections not averted. We conducted the same counterfactual calculations in two other hypothetical scenarios: one for policy changes that resulted in a conservative low reduction in PrEP coverage (2 per 100 decrease), and one for policy changes that resulted in a potential high reduction in PrEP coverage (10 per 100 decrease).

Cost analyses were carried out based on our counterfactual infection case predictions. To estimate net costs due to decreases in PrEP investment over 10 years, we applied current estimates of HIV-related treatment costs to estimate costs incurred because of HIV infections not averted by PrEP. To calculate net incremental costs, we subtracted the savings realized from providing PrEP to fewer people. We used undiscounted annual HIV-related medical costs per person living with HIV^21^ to estimate the increase in 10-year HIV medical costs associated with unprevented HIV infections. Similarly, we used lifetime HIV-related medical costs per person living with HIV^21^ ($420,285 2019 US$, discounted at 3%; or $1,079,999 (2019 US$), undiscounted) to estimate the increase in lifetime HIV medical costs associated with unprevented infections. Discounted analyses indicate the present value, reflecting the time value of money; undiscounted analyses represent the total projected cash flows associated with the scenario. The corresponding net costs over a decade were calculated by subtracting estimated medical costs for PrEP for people who were assumed to not be taking PrEP under an adverse policy scenario.

All analyses were conducted using SAS software version 9.4 (SAS Institute, Cary, North Carolina) and the Joinpoint Regression Program, version 5.0.2 (National Cancer Institute). The Poisson log-linear models were carried out using the GIMMIX procedure in SAS.

## Results

Assuming that negative changes in PrEP programs reduced PrEP coverage yearly by 3.3 per 100 over the next decade (e.g., impacts of discontinuing interventions to increase awareness of PrEP, increasing out of pocket costs of PrEP, rolling back the expansion of health coverage and drug assistance programs, and/or decreasing retention in PrEP care), HIV diagnosis rates were predicted to increase by an average 2.3% annually. Even this modest decrease in PrEP coverage would be predicted to erase all the reductions in HIV transmissions achieved over the past decade. In this scenario, 8,618 new infections would fail to be averted in a decade because of lowered PrEP uptake; the estimated lifetime medical costs of these unaverted infections would be $3.6 billion (discounted) and $9.3 billion (undiscounted). The net costs over a 10-year period in this scenario would be $1.4 billion (discounted) and $1.9 billion (undiscounted). We also evaluated a scenario of larger cuts to PrEP programs, estimates of yearly decrease in PrEP coverage of 10 per 100. This scenario resulted in 26,873 HIV infections that failed to be averted in a decade with correspondingly higher lifetime medical costs--$11.3 billion (discounted) and $29.0 billion (undiscounted) (Table 1).

**Table 1.**
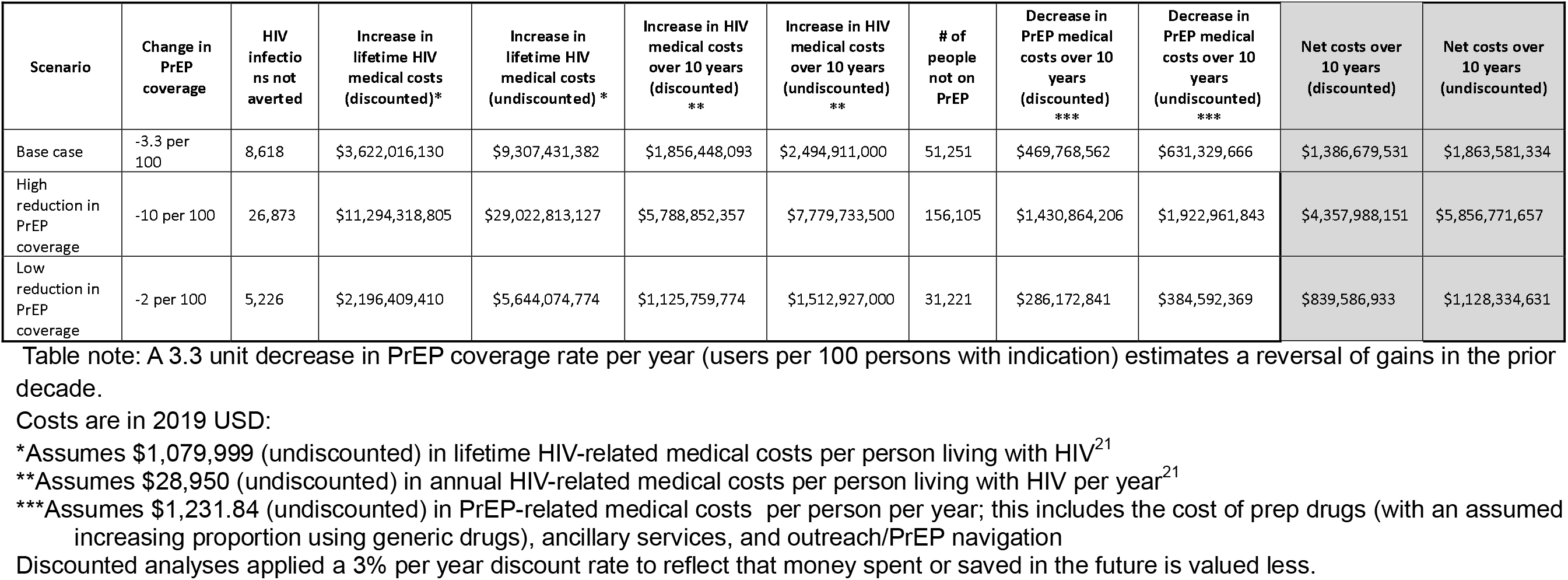
Estimated effects of reduced PrEP coverage on HIV diagnoses and costs over 10 years, United States.

| Scenario | Change in PrEP coverage | HIV infections not averted | Increase in lifetime HIV medical costs (discounted)* | Increase in lifetime HIV medical costs (undiscounted) * | Increase in HIV medical costs over 10 years (discounted) ** | Increase in HIV medical costs over 10 years (undiscounted) ** | # of people not on PrEP | Decrease in PrEP medical costs over 10 years (discounted) *** | Decrease in PrEP medical costs over 10 years (undiscounted) *** | Net costs over 10 years (discounted) | Net costs over 10 years (undiscounted) |
| --- | --- | --- | --- | --- | --- | --- | --- | --- | --- | --- | --- |
| Base case | -3.3 per 100 | 8,618 | \$3,622,016,130 | \$9,307,431,382 | \$1,856,448,093 | \$2,494,911,000 | 51,251 | \$469,768,562 | \$631,329,666 | \$1,386,679,531 | \$1,863,581,334 |
| High reduction in PrEP coverage | -10 per 100 | 26,873 | \$11,294,318,805 | \$29,022,813,127 | \$5,788,852,357 | \$7,779,733,500 | 156,105 | \$1,430,864,206 | \$1,922,961,843 | \$4,357,988,151 | \$5,856,771,657 |
| Low reduction in PrEP coverage | -2 per 100 | 5,226 | \$2,196,409,410 | \$5,644,074,774 | \$1,125,759,774 | \$1,512,927,000 | 31,221 | \$286,172,841 | \$384,592,369 | \$839,586,933 | \$1,128,334,631 |

Table note: A 3.3 unit decrease in PrEP coverage rate per year (users per 100 persons with indication) estimates a reversal of gains in the prior decade.
Costs are in 2019 USD:
\*Assumes \$1,079,999 (undiscounted) in lifetime HIV-related medical costs per person living with HIV<sup>21</sup>
\*\*Assumes \$28,950 (undiscounted) in annual HIV-related medical costs per person living with HIV per year<sup>21</sup>
Discounted analyses applied a 3% per year discount rate to reflect that money spent or saved in the future is valued less.

## Discussion

Nearly 38,000 new HIV infections occurred in the United States and territories in 2022.^22^ There is currently no vaccine or cure that is imminent, so PrEP is the necessary mainstay of public health programs. HIV prevention via PrEP improves the health of Americans and can save money in terms of medical costs from the payer perspective.^19^ Moreover, though not considered here, cost estimates from the societal perspective would likely show even more cost savings, because most HIV infections occur in younger people.^23^

According to our analyses, any program or policy changes that reverse the gains in PrEP coverage that have been made over the past decade will result in increased new HIV infections and substantial increased costs to the healthcare system and society. Conversely, by maintaining or expanding current effective programs, the United States could avert billions of dollars in medical costs associated with infections that would not be averted following disinvestment in HIV prevention. We focus here on possible threats to maintaining PrEP coverage; conversely, programs that increase investments in PrEP programs offer the opportunity to increase PrEP coverage, which has been reported to be cost-saving in multiple prior analyses.^18,24,25^

Our cost estimates are conservative, in that the costs included in our calculations only address the lifetime medical costs of those people who are estimated to acquire new HIV infections because of less PrEP coverage. We do not include cost of secondary infections that are estimated to occur from individuals acquiring HIV in our model in the interval between their HIV infection and their diagnosis and effective treatment^26^. Including such onward transmissions would significantly increase our estimates of costs associated with failing to sustain current support for PrEP programs. Our analyses assume that no other programmatic changes will be made – for example, reductions to the Ryan White Care Act or other programs that support people engaged in HIV care. If such other changes occur, our data would reflect a minimum estimate of impacts on new infections.

Our analysis has some important limitations. We rely on a quasi-experimental approach to estimate the associations between levels of PrEP coverage and subsequent HIV infections, and we assume that decreasing PrEP coverage will result in mathematically inverse effects on new HIV diagnoses. Systems of PrEP delivery are complicated, and interventions and many other factors can influence trends in new HIV diagnoses, but the observed relationships between population levels of PrEP coverage and new HIV diagnoses are based on real world data and are robust. Although ecological, these relationships meet many of the criteria for causality^27^, including biological plausibility, dose response relationships (biological gradient), temporality, coherence, and consistency. Further, there are other factors that shape the actual impact of PrEP, and for which we did not account in our analyses. For example, we did not evaluate the impact of the extent to which those at highest risk for infection are the people receiving PrEP, adherence, viral resistance to active components of PrEP medications, or the adjunctive use of other prevention modalities. Although these considerations are important in considering the risk of individuals, at the level of populations, we believe that the association of the extent of PrEP coverage and population-level new HIV infections is distinguishable. This is characteristic of the strengths and limitations of analyses of real-world evidence.

HIV prevention using PrEP works at the individual level, and a decade of research has documented that PrEP coverage at the population level is associated with reduced new HIV infections and diagnoses. Using real-world data about experiences with PrEP in the United States, we estimated that decreasing coverage of PrEP would have substantial negative impacts on new HIV infections. Such changes in coverage would be the expected results of disinvestment in HIV prevention activities or policy changes that discourage PrEP use by increasing out of pocket costs for PrEP, or by increasing barriers to HIV screening, which is the gateway to PrEP services.

Consideration of policies that would reduce access to PrEP could not come at a worse moment. After sustained national investments, we now have not only a daily pill option for PrEP^2^, but injectable PrEP that can be administered every two months.^28^ Injectable PrEP that only requires two shots a year is being considered for approval by the US FDA^29^, and there is potential for an annual shot.^30^ Injectable cabotegravir shows strong evidence for HIV prevention in women.^31^ Each of these advancements makes it easier to achieve greater population-level coverage of PrEP and associated reductions in HIV and to optimize the payoff from sustained national investments in HIV prevention and care. Disinvestment in PrEP programs would slow the current progress in reducing new infections; disinvestment in PrEP research structures would also destabilize a present (and desperately needed) acceleration in HIV prevention technologies. We are in a time of increasing PrEP choices and declining new HIV diagnoses in the United States; any obstacles to sustained HIV prevention services, including access to HIV screening and PrEP, will burden our country by increasing health and financial costs.

## Data Availability

All data used in these analyses are publicly available through AIDSVu.org and CDCAtlasPlus

https://www.cdc.gov/nchhstp/about/atlasplus.html

https://aidsvu.org/

